# Preventing psychological symptoms among cancer survivors through a digital mindfulness psychoeducation program: Protocol of a randomized controlled trial

**DOI:** 10.1101/2023.01.05.23284230

**Authors:** Piyanee Klainin-Yobas, Kanokwan Hounsri, Wee Joo Chng, Neo Kim Emily Ang, Yong-Shian Shawn Goh

## Abstract

**Background:** Cancer survivors experience challenges, that may affect their psychological well-being. Technology-based, mindfulness-based interventions have been offered to cancer survivors; however, contents do not sufficiently cover issues related to cancer and its treatments. This study protocol presents a three-arm randomized controlled trial (RCT) that aims to examine the effectiveness of a digital mindfulness psychoeducation programme (Digital-MindCAN programme) on psychological symptoms among cancer survivors. This program contains knowledge linking to cancer-related matters, mindfulness principles and mindfulness practice, which will be delivered real-time using a videotelephony software.

**Methods:** Eligible cancer survivors will include adults who completed curative treatments from two weeks to two years. They will be randomly assigned to one of the three groups: Digital-MindCAN programme with standard care, Palouse Mindfulness programme with standard care, and a wait-list control group. A convenience sample will be recruited from a tertiary hospital in Singapore. A target sample size will be 150 participants, with 50 participants in each group. Primary outcomes encompass objective stress and subjective stress. Secondary outcomes comprise psychological well-being, perceived relaxation, mindfulness, resilience, depression, and anxiety. Self-administered questionnaires and physiological measures will be used to collect participants’ responses. Focus group interviews will be conducted for intervention groups after the end of the eighth session. Quantitative data will be analyzed by descriptive statistics, analysis of covariance and repeated measures analysis of variance. Qualitative findings will be analyzed using a realist evaluation method.

**Discussion:** This RCT will be the first to test the effectiveness of the technology-based, mindfulness-based intervention on cancer survivors in Singapore. Positive findings will add knowledge and inform clinical practice. Specifically, the Digital-MindCAN intervention may be offered as part of standard care for cancer survivors. Future research can be implemented and further tested the program in other healthcare institutions.

**Trial registration:** This study has been registered with ISCTN Clinical Trial Registry (Trial NO. ISRCTN10756933).

## Introduction

Cancer is a major global health problem. Global cancer statistics in 2020 indicated that there were approximately 10.0 million cancer deaths and 19.3 million new cases of cancer worldwide. In particular, breast cancer is the most diagnosed type, which account for 2.26 million of all cancer diagnoses [1]. As in Singapore, the incidence and prevalence of cancer continue to increase, and thus it has the highest age-standardized rate of cancer in Southeast Asia [2]. During the period of 2015 - 2019, 78,204 cancer cases were reported in Singapore with 49% females and 51% males [3]. Most cancer survivors experienced physical and psychological impacts that interfered with their daily lives. Such impacts included physical limitations, mood swings, fear of recurrence, difficulty in returning to work, and financial obligations [4].

Depression, anxiety, and stress are frequent symptoms in patients with cancer. Depression is characterized by persistently depressed mood, incapacity to show interest in pleasurable activities, cognitive impairment, feelings of worthlessness, deficits in attention, and suicidal thoughts [5]. Stress refers to acute or chronic exposure to the stressor and the response, which increases catecholamines and cortisol mediator in the bloodstream [6]. Most cancer survivors are particularly more susceptible to depression and stress than the general population [7]. Anxiety is manifested by excessive worries about the progression of the disease, and the difficulty to control physical symptoms including restlessness, fatigue, concentration struggle, impatience, sleep disturbances, and muscle tension [5]. Patients with cancer, especially younger ones, were more anxious in comparison with the general population [8].

Resilience is a crucial personal factor for cancer survivors. Resilience may provide a protection against the devastating consequences of stress by lessening or absorbing the shock of a cancer diagnosis, the impact of disgusted events, and related life changes [9]. Cancer patients with high resilience experienced lower psychological distress and physically more active [10]. At the same time, cancer patients with a higher level of resilience were reported to have improved quality of life, less depression and anxiety, and better function [11].

Psychological well-being refers to an attempt to improve and fulfill potential, which is related to having a purpose in life, coping with challenges, and making a certain effort to achieve goals [12]. The experience of cancer can be damaging to the social life, work, and functioning and psychological well-being of individuals [13]. A systematic review revealed that mindfulness-based interventions enhance psychological well-being among breast cancer survivors with a small effect size (SMD=0.39) [14].

Mindfulness intervention has the potential to influence cancer survivors’ physical and psychological outcomes. Mindfulness is defined as awareness of thoughts, emotions, and sensations, with an emphasis on cultivating a nonjudgmental manner to the present moment [15]. The most frequently used interventions include mindfulness-based cancer recovery (MBCR), mindfulness-Based stress reduction (MBSR), and mindfulness-based cognitive therapy (MBCT). MBCR is a group-based mindfulness training that teaches people living with cancer mindfulness meditation and movement practices [16]. In addition, MBSR and MBCT usually begin with a simple body scan, mindfulness of breath, and movement practices, which have benefits on mental health outcomes [17].

In additional to a face-to-face, traditional approach, mindfulness-based interventions have been delivered via technology-based platforms such as online video, video conference, audio compact disk, MP3 player, telephone, mobile application, and internet [18]. Such platforms are perceived to be cost-effective and accessible for cancer survivors, especially during the COVID-19 pandemic. A systematic review of 18 studies suggested that eHealth mindfulness-based interventions improved anxiety, depression, mindfulness level, and quality of life among cancer survivors [18]. Furthermore, the review suggested that a intervention duration should range from four to eight weeks and various techniques can be utilized such as lectures and online group discussions. To enhance awareness, mindfulness practice can be performed such as body scan practice, moving meditation, hatha yoga and loving-kindness meditations [18].

Although there are different paradigms of mindfulness-based interventions, they show positive influences on improving anxiety, depression, and quality of life. They also reduce fatigue, stress, and posttraumatic growth among adult cancer patients and survivors [19]. However, there are a few gaps in the existing literature of mindfulness-based interventions. Firstly, most mindfulness-based interventions focus on mindfulness practice but do not sufficiently offer knowledge specific for cancer survivors. Secondly, while most studies have examined how a mindfulness-based intervention can minimise psychological symptoms, it is unclear whether such interventions can prevent these symptoms. Thirdly, it is inconclusive that a mindfulness-based intervention can enhance psychological well-being and resilience. Moreover, previous studies have primarily focused on quantitative data and overlooked qualitative information. Finally, most research has relied on the use of self-reported questionnaires and overlook objective measures to assess stress levels.

### The current Study

To minimize the lacuna in the literature, the digital mindfulness-based psychoeducation for cancer survivors (Digital-MindCAN Program) was developed with contents tailoring specifically for cancer survivors. The Digital-MindCAN program will be offered in an interactive group format and will be delivered real-time by the researchers using a videotelephony platform. This paper will present a study protocol that aims to examine the effectiveness of the Digital-MindCAN Program in comparison with an active control and wait-list control group. Schedule of enrolment, intervention and assessment is shown in Fig 1

**Fig 1.**
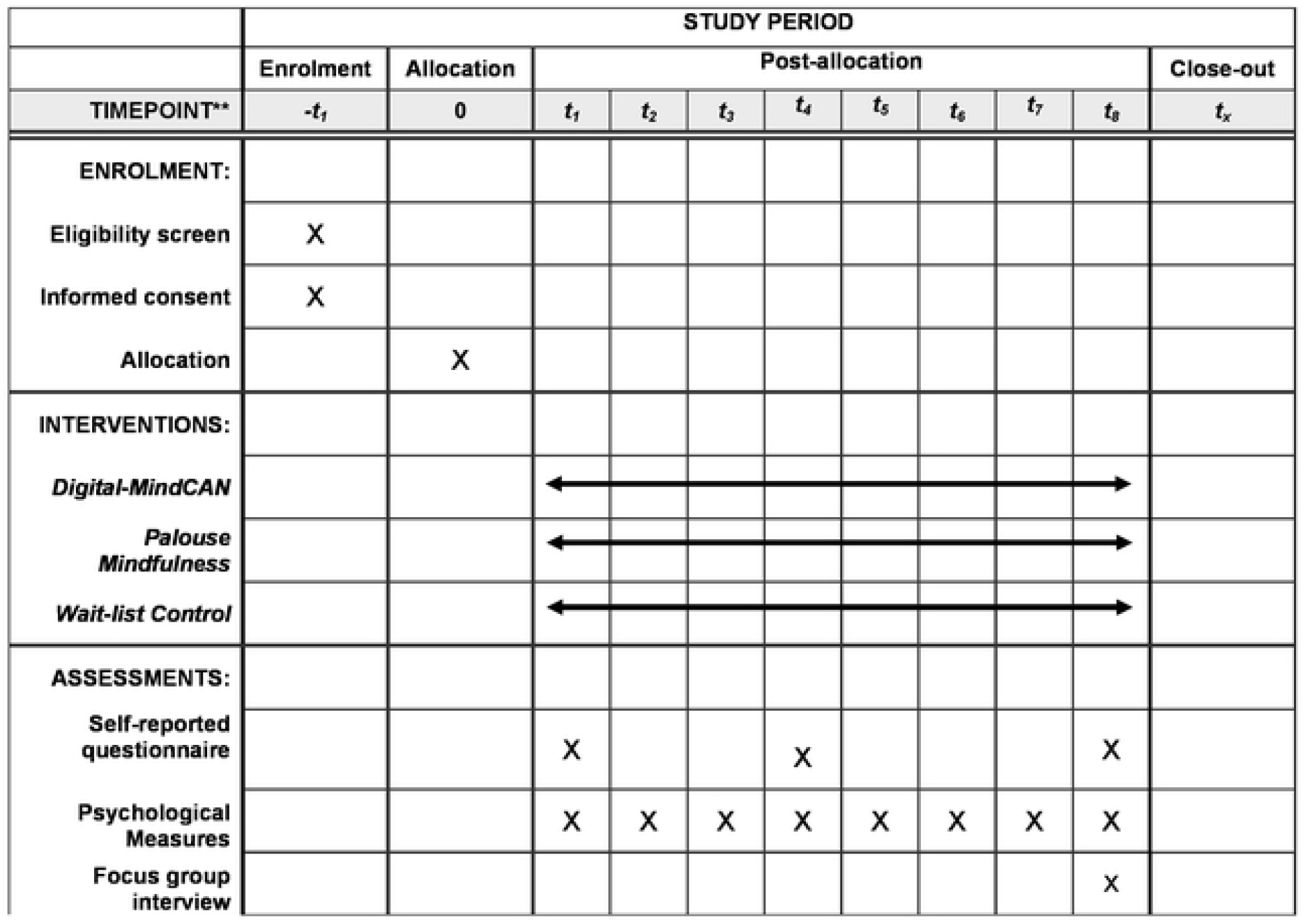
Schedule of enrolment, interventions, and assessments. -Self-reported questionnaire includes Psychological well-being, Mindfulness, Resilience, Subjective Stress, Depression, Anxiety -Psychological measures include Stress Thermometer, Oxygen Saturation, and Heart Rate

Research questions are:

1. In comparison to wait-list control, will participants in the Digital-MindCAN group report significantly greater improvement in objective stress, subjective stress, depression, anxiety, perceived relaxation, psychological well-being, mindfulness, and resilience?
2. Will the Digital-MindCAN group be non-inferior than the active control group (receiving Palouse Mindfulness) in improving objective stress, subjective stress, depression, anxiety, perceived relaxation, psychological well-being, mindfulness, and resilience?
3. What is participants’ perception towards the Digital-MindCAN or Palouse Mindfulness intervention?

### Theoretical Framework

The psychological well-being (PWB) promotion model (Fig 2) provides a theoretical framework to guide this study. This model was created by the researchers according to the Neuman’s system theory [20] and Ryff’s psychological well-being [21]. It is postulated that stress has a negative impact on individuals’ PWB (cancer survivors in this study). However, prevention interventions (primary, secondary and tertiary prevention) will help the individual restore their PWB. The Digital-MindCAN program represents the secondary prevention intervention, which help cancer survivors deal with cancer-related stressors, prevent psychological symptoms and resume their PWB.

**Fig 2.**
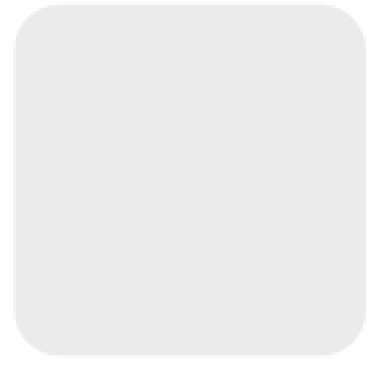
The Psychological Well-being Promotion Model.

## Materials and methods

This study follows the CONSORT2010 guidelines for reporting parallel group randomized trials [22]. We also registered this RCT with ISCTN Clinical Trial Registry (ISRCTN10756933). Furthermore, this study has been approved by NHG Domain Specific Review Board, Singapore [NHG DSRB 2021/00083].

### Study design and setting

A three-arm, parallel, randomized controlled trial will be conducted and participants will be randomly assigned with a 1:1:1 ratio to either: a) an intervention group receiving the Digital-MindCAN programme and standard care, b) active control group receiving the Palouse Mindfulness intervention and standard care, or c) wait-listed control group. Participants will be recruited from a cancer center at a tertiary hospital in Singapore. The cancer center utilizes a multidisciplinary and holistic approach; and offers one-stop services including prevention, diagnostic, and wide ranges of evidence-based treatments and advanced technology to people with cancer.

Blinding of participants will be challenging as participants in the control groups will be aware of their allocation. Nonetheless, people in the intervention groups will not know which group is the intervention or active control. Blinding of outcome accessors will also be difficult. However, given that some variables will be measured by physiological measures (such as pulse oximeter and stress thermometer), detection bias may be minimized.

### Participants

To be eligible, participants must be: 1) adults of any gender aged 21 to 65 years with a diagnosis of any type of cancer stage 0 – III by their attending physician, and b) those who completed all cancer and related treatments (such as chemotherapy, surgery, and radiotherapy) as prescribed by their attending physician between 2 weeks to 2 years after the diagnosis, with the exception of hormonal therapy [23]. The exclusion criteria for this study are individuals who: 1) are pregnant, 2) had severe mental disorders (e.g., schizophrenia, mood disorders, bipolar disorder, and substance-related disorders), and 3) had severe medical conditions requiring hospitalisation. The mental health and medical conditions should be diagnosed by physicians. The conditions are perceived to link with additional stressors, which might confound our research findings.

### Sample size calculation

An adequate sample size is crucial to detect statistically significant and clinically important differences in the phenomena of interest [24]. In line with this, power analysis was performed using G*Power v3.1.9.2 to calculate an adequate sample size for this study. Using an effect size of 0.35 obtained from a previous systematic review examining the effect of mindfulness interventions on stress [19], three groups, power of 80%, and two-sided 5% significance level, a total sample size will be at least 142. This number was rounded up a total sample size of 150, with 50 participants in each group.

### Randomization Process

This randomization process encompasses three steps: Sequence generation, allocation concealment, and implementation [22]. First, a *random allocation sequence* will be generated by the principal investigator (PI), who will not involve in recruitment and assessment process. Specifically, a serial number of 1-150 will be generated and randomly assigned into three groups using Microsoft Excel software, forming a random allocation list. Secondly, *allocation concealment* is an important mechanism to prevent recruiters’ pre-awareness of the treatment allocation, which might lead to selection bias during the recruitment process [22]. Therefore, the PI will conceal the allocation list and release it to the recruiters after the potential participants sign the written consent form and complete all relevant procedures. During the *implementation*, participants who are enrolled to this study, will receive a unique identification number (ID) according to the order of their enrollment. Subsequently, they will be allocated to the group containing pre-assigned number that match their ID.

### Recruitment Procedure

The researchers will liaise with attending healthcare professionals (such as nurses and physicians) in identifying potential participants. Once identified, the researchers will approach eligible clients, offer them a brochure and participant information sheet, explain the aims of the study and invite them to take part in the study. They will be given time to ask any questions they may have regarding the study. If the clients need more time to consider, they can contact the researchers later at their own convenience. If the clients are eligible and wish to sign up for the study, the researchers will invite them to a private room provided by the clinic. Should the individuals agree to participate in the study, a written informed consent will be signed. Subsequently, the clients will be informed about their group allocation. Those who are assigned to the Digital-MindCAN and Palouse mindfulness groups will be informed about the intervention schedule, venue, and data collection. After those two groups complete the interventions and data collections, participants in the wait-list control will be offered to attend either Digital-MindCAN or Palouse mindfulness intervention. However, data during the interventions will not be collected for testing the interventions. Each participant will receive an honorarium of 100 Singapore Dollar (about 74 US Dollar) for participating in this study.

### Intervention: Digital-MindCAN programme

The Digital-MindCAN programme aims to help cancer survivors learn to manage stress and regulate unpleasant emotions. The contents of the programme entail information specific for cancer survivors, such as stress related to cancer symptoms, side effects of cancer treatments, and management of thoughts and emotions commonly occurring among cancer survivors [25]. The weekly, group-based, Digital-MindCAN programme comprises eight sessions with two components: education and mindfulness practice. Each session lasts 90 minutes. Participants will be asked to perform mindfulness practices at home for 10 to 30 minutes a day and record their practice in a Mindfulness Practice Diary (Fig 3).

**Fig 3.**
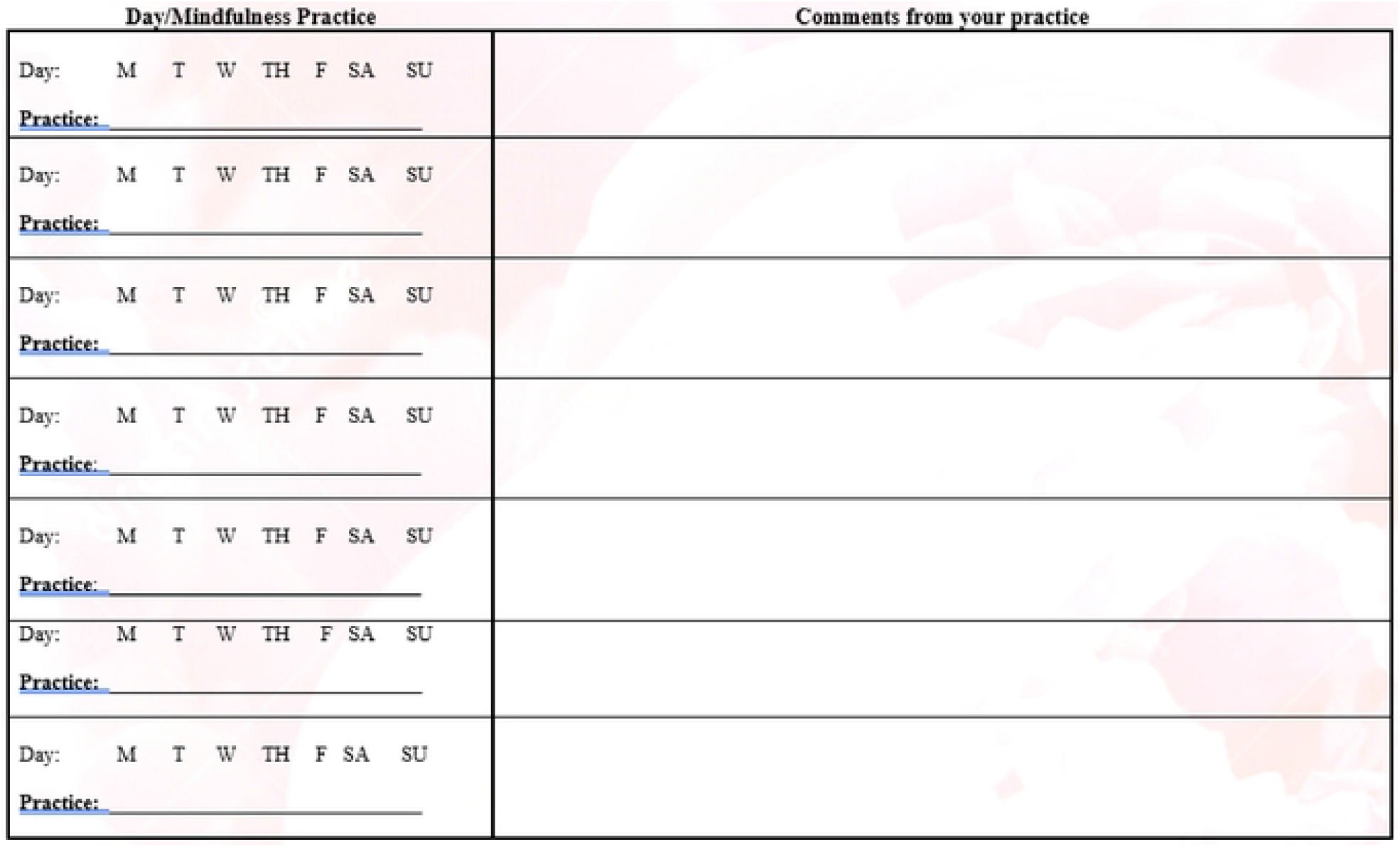
Mindfulness Practice Diary.

The education components will cover the following topics: 1) The world of mindfulness, 2) Mindful management of stress, 3) The Field of awareness: Mindful body sensations, 4) Mindful emotions: Calmness and composure, 5) Mindfulness: The powerful mind: 6) Mindful communications, 7) Loving-kindness and compassion, and 8) Mindful living: Building your mindful life style. Concerning the mindfulness practice, participants will learn to perform mindful breathing, STOP breathing practice, body scan practice, the five-step PAIN management process, mindfulness of positive emotions, letting-go meditation, mindful sitting, and loving kindness mediation. Furthermore, all participants will receive a MindCAN education booklet so that they can review the materials at their convenience. Additionally, a mobile messaging platform will be created to facilitate ongoing communications between the researchers and participants outside the intervention sessions.

### Active control intervention: Palouse Mindfulness programme

The Palouse Mindfulness intervention [26] is an internet-based MBSR programme developed and modified based on Kabat-Zinn’s MBSR [15]. It entails eight sessions, and each session lasts 90 minutes. The Palouse Mindfulness is a general stress reduction programme without contents specific for cancer. Each week, participants will learn about mindfulness principles and perform mindfulness practice. Weekly topics include: 1) Simple awareness, 2) Attention and the brain, 3) Dealing with thoughts, 4) Stress: Responding and reacting, 5) Dealing with difficult emotions or physical pain, 6) Mindfulness and communication, 7) Mindfulness and compassion, and 8) Conclusion: Developing a practice of your own. Regarding the mindfulness practice, participants will perform sitting meditation, body scan, mountain meditation, lake meditation, loving kindness and Rain meditation. Furthermore, participants are to perform mindfulness practices at home for 10 to 30 minutes daily and record their practice in a Mindfulness Practice Diary (Fig 2). All participants will also receive a Palouse Mindfulness booklet and will be part of a mobile messaging group.

### Wait-list control group

Participants in the wait-listed control group will not attend any intervention during the period that the Digital-MindCAN programme and Palouse Mindfulness are implemented. They are to provide information through self-reported questionnaires and physiological measurements on a certain day of each week for eight weeks. Such data collections will be scheduled based on their convenience. After the interventions and data collection for intervention groups are completed, the wait-listed participants can request to attend any intervention (either the Digital-MindCAN or Palouse mindfulness) if they wish. The researchers will then organise the interventions at their convenience. The interventions provided to the wait-listed participants will not be part of the study; therefore, no additional data will be collected.

### Measures

Data will be collected by self-reported questionnaires, physiological measurements, and focus group interviews (Table 1). Primary outcomes are objective stress and subjective stress. Secondary outcomes comprise psychological well-being, perceived relaxation, mindfulness, resilience, depression, and anxiety. psychological well-being, subjective stress, mindfulness, and resilience, as self-reported questionnaires, will be assessed three times: at baseline, after the fourth session, and after the eighth session. To capture immediate changes, objective stress and perceived relaxation will be measured before and after each intervention sessions. Accordingly, 16 sets of data will be collected. Furthermore, focus group interviews will be conducted after the eighth session.

**Table 1.** Assessments for Intervention and Control Groups

| Palouse Mindfulness<br>MindCAN groups | Intervention Sessions (Week) |  |  |  |  |  |  |  |  |  |  |  |  |  |  |  |
| --- | --- | --- | --- | --- | --- | --- | --- | --- | --- | --- | --- | --- | --- | --- | --- | --- |
| Assessment | Baseline |  | 2 |  | 3 |  | 4 |  | 5 |  | 6 |  | 7 |  | 8 |  |
|  | Pre | Post | Pre | Post | Pre | Post | Pre | Post | Pre | Post | Pre | Post | Pre | Post | Pre | Post |
| Objective Stress<br>- Stress Thermometer<br>- Oxygen Saturation<br>- Heart Rate | ✓ | ✓ | ✓ | ✓ | ✓ | ✓ | ✓ | ✓ | ✓ | ✓ | ✓ | ✓ | ✓ | ✓ | ✓ | ✓ |
| Perceived Relaxation |  | ✓ |  |  |  |  |  |  |  | ✓ |  |  |  |  |  | ✓ |
| Psychological Outcomes<br>(Psychological well-being,<br>Mindfulness, Resilience,<br>Subjective Stress,<br>Depression, Anxiety) |  | ✓ |  |  |  |  |  |  |  | ✓ |  |  |  |  |  | ✓ |
| Focus Group Interview |  |  |  |  |  |  |  |  |  |  |  |  |  |  |  | ✓ |
| Control Group | Research Period (Week) |  |  |  |  |  |  |  |  |  |  |  |  |  |  |  |
| Assessment | Baseline |  | 2 |  | 3 |  | 4 |  | 5 |  | 6 |  | 7 |  | 8 |  |
|  | 1.1 | 1.2 | 2.1 | 2.2 | 3.1 | 3.2 | 4.1 | 4.2 | 5.1 | 5.2 | 6.1 | 6.2 | 7.1 | 7.2 | 8.1 | 8.2 |
| Objective Stress<br>(Stress Thermometer,<br>Oxygen Saturation, Heart<br>Rate) | ✓ | ✓ | ✓ | ✓ | ✓ | ✓ | ✓ | ✓ | ✓ | ✓ | ✓ | ✓ | ✓ | ✓ | ✓ | ✓ |
| Perceived Relaxation |  | ✓ |  |  |  |  |  |  |  | ✓ |  |  |  |  |  | ✓ |
| Psychological Outcomes<br>(Psychological well-being,<br>Mindfulness, Resilience,<br>Subjective Stress,<br>Depression, Anxiety) |  | ✓ |  |  |  |  |  |  |  | ✓ |  |  |  |  |  | ✓ |

**Objective stress** will be measured by peripheral skin temperature, heart rate, and oxygen saturation (SpO_2_) variability. Peripheral skin temperature will be measured by a stress thermometer [27]. All respondents will be asked to place a temperature sensor between their thumb and finger index for one minute. The researcher will provide instructions on how to use the equipment, record the values, and assist the respondents during the measurement to ascertain procedural standardization. In addition, the room temperature within the range of 68°F to 77°F (20°C - 25°C) will be set to ensure environmental consistency across all intervention sessions. The stress thermometer provides a measurement range from 58 °F to 158 °F (14.4°C to 70°C), with lower scores suggesting higher objective stress. A previous study by reported that an interclass correlation coefficient across sessions conducted in cancer survivors was 0.80, suggesting good reliability [25].

Heart rate and oxygen saturation (SpO_2)_ variability will be measured by the LAICA EA1006 Pulse Oximeter. The oxygen saturation level less than 85% represents “several hypoxic” whereas 95%-100% are found normal in healthy individuals [28]. In terms of heart rate, a normal resting heart rate for adults ranges from 60 to 100 beats per minute [29]. A higher heart rate represents a higher stress level in this study [30].

**Subjective stress, depression, and anxiety** will be measured by the 21-item Depression Anxiety and Stress scale (DASS-21) [31]. The DASS-21 contains three subscales: depression, anxiety and depression. Each subscale has seven items with response categories ranging from 0 (did not apply to me at all) to 4 (applied to me most of the time). Total scores for each subscale range from 0 to 28, with higher scores indicating the symptom. Cronbach’s alpha of the scale in a previous study was 0.85 [25].

**Psychological well-being** will be measured using the 18-item Psychological Well-being Scale [21]. Responses to each question range from 1 (strongly disagree) to 6 (strongly agree). Total scores range from 18 to 108, with higher scores indicating greater well-being. Cronbach’s alpha of the scale among cancer survivors was 0.78 [25].

**Perceived relaxation** will be assessed by the perceived relaxation scale [32]. The visual analogue scale comprises a horizontal line with numbers to convey the degree of relaxation. The patients will rate their level of relaxation using the scale from 0 (very tense) to 10 (very relaxed). Higher scores reflect higher levels of perceived relaxation. Reliability of the scale was established in a previous study among cancer survivors as an interclass correlation coefficient across sessions was 0.80 [25].

**Mindfulness** will be measured using the 15-item Mindful Attention Awareness Scale (MAAS) [33]. Participants will rate on six-point categories ranging from 1 (almost always) to 6 (almost never). Total scores range from 1 to 90, with higher scores signifying greater levels of mindfulness. Cronbach’s alpha of the scale among cancer survivors as 0.92 [25].

**Resilience** will be assessed by the 10-item Connor-Davidson Resilience Scale [34]. Each item is rated on a 5-point scale ranging from 0 (not true at all) to 4 (true all the time). Total scores range from 0 to 40, with higher scores indicating higher levels of resilience. In a previous study on cancer survivors, Cronbach’s alpha of the scale was 0.93 suggesting excellent reliability [25].

**Perception towards the interventions** (**MindCAN or Palouse Mindfulness)** will be assessed by focus group interviews, which will be conducted at the end of the eight session. Focus group interview is a technique to elicit in-depth information on the phenomena of interest [35]. The nature of group dynamic and interactions among group member help generate deeper and richer information than that from individual interview [35]. The interviews will be conducted by the PI or by co-investigators who have been trained to conduct such interviews. Focus group interviews will be conducted separately for the Digital-MindCAN and Palouse Mindfulness groups. Each interview group will entail four to eight participants from the same cohort. The interviews will last approximately one hour and they will be audio-recorded. An interview guide will be used for both MindCAN and Palouse Mindfulness groups. Examples of the interview questions are: 1) “What are your perceptions of the MindCAN program (Palouse Mindfulness)?, 2) What do you think about the contents of the programs?, 3) What are your experiences with the mindfulness practice during the session and at home?, and 4) What would be your suggestions to improve the program?

**Treatment fidelity** will be documented by group facilitators. Specifically, participants’ attendance rate will be recorded and monitored during the intervention periods. The facilitators will also keep a weekly log to document important discussions, activities and matter arising during the group process. Additionally, group members (Digital-MindCAN and Palouse Mindfulness groups) will be encouraged to keep the Mindfulness Practice Diary (Table 2), containing the record of their home practice and feelings during the practice.

**Personal information** will include age, gender, nationality, ethnicity, religion, employment, monthly gross household income, and satisfaction level of the income. Health information will consist of medical diagnosis, the time received the diagnosis, and received treatments.

### Data Analyses

All research data will be entered into the IBM SPSS software version 28. To ascertain the accuracy of data entry, one researcher will enter the data while another researcher will check the entered data against the corresponding questionnaire. Data cleansing (using Frequency and Crosstabulation) will be operated on each variable to ensure that there are no values out of range. Intent-to-treat analysis will be used to ensure that all data from randomized participants are included [36]. Missing data will be handled with a multiple imputation method [37]. Baseline comparability will be performed whereby characteristics and study variables of participants will be tabulated for each group [36]. The tabular presentation will allow readers to judge the success of the randomization [36]. Then, the study variables will be summarized into descriptive statistics (mean, standard deviation, and frequency).

Mean differences in subjective stress, depression, anxiety, mindfulness, resilience, and psychological well-being across three assessment points and three groups will be tested by analysis of covariance (ANCOVA). Covariate adjustment will be performed whereby a baseline score for each variable will serve as a covariate [36]. Such covariate adjustment can prevent the effect of covariate imbalance, the presence of strong correlation between the baseline score and study outcomes [36]. Mean differences in skin temperature, heart rate, SPO_2_ and perceived relaxation will be tested by repeated measures analysis of variance. Partial eta square (η_p_^2^) will be used as an effect size measure to determine the effect of the Digital-MindCAN program on study variables. The η_p_^2^ values of 0.01, 0.06, and 0.14 will represent a small, medium, and large effect size respectively [24].

The qualitative data on focus group interviews will be analyzed by a realist analysis, which aims to obtain in-depth understanding of which contexts and mechanisms of the mindfulness interventions can contribute to study outcomes [38]. Unlike traditional qualitative data analyses, the realist approach takes into accounts both inductively-derived themes (from study participants) and pre-existing theoretical-deductive conceptualizations (from the literature). The realist analytical approach comprises five steps: 1) Transcriptions and Indexing, 2) Interpretation and Theorization, 3) Contrastive demi-regularities, 4) Retroduction of generative mechanisms, and 5) Concretization [38]. First, *Transcription* will involve taking and reviewing all information from focus group discussions while *Indexing* will be performed by assigning non-exclusive codes on the raw materials and maintaining points/issues originally raised by the participants.

Secondly, *Interpretation*, an abstraction process, requires an interpretative understanding of the indexed data (codes) and inductively generating conceptual themes. Possible explanations of the mechanisms underlying the mindfulness interventions can be produced at this process [38]. During the *Theorization*, the researchers will establish theoretically-deduced themes and mechanisms of the interventions that drawn from the existing literature. Thirdly, *Contrastive demi-regularities* are concerned with identifying contradictory findings. Fourthly, *Retroduction* will synthesize all information (including inductively-derived themes and deductively-related theories). Finally, *Concretization* will involve applying and/or re-contextualizing all the retroduced generative structures to elaborate participants’ perception of the mindfulness interventions.

### Ethical considerations

This study received ethics approval from the Domain Specific Review Board (DSRB), National Healthcare Group, Singapore [NHG DSRB Ref: 2021/00083]. All participants will complete a written consent form, which include statements linking with such issues like confidentiality, voluntary participation, safeguarding and study withdrawal. Each raw questionnaire will be anonymized and will be assigned a unique identification number. The interview transcriptions will use pseudonyms. No identifiable information will be used in any document, research report, or publications. If the interventions or questionnaires are perceived to cause harm to participants, they will be advised to discontinue the study and all data will be destroyed. All research data will be kept in the PI locked office and only the research team will have access to the data.

## Discussion

This RCT aims to generate empirical evidence to support that the eight-week, group-based, Digital-MindCAN program is: a) non-inferior than the original mindfulness-based stress reduction intervention (Palouse Mindfulness) and b) superior to the wait-listed control group in improving psychological outcomes. Positive findings from this study will add knowledge to the literature concerning the usefulness and effective of the Digital-MindCAN program among cancer survivors in Singapore. The results may inform clinical practice in that the program can be offered as an alternative service for cancer survivors to help them manage stress and enhance psychological well-being. Future research may explore the possibility of using other digital platforms including mobile application, virtual reality, augmented reality and artificial intelligence.

We anticipate some challenges that might take place during the research process. One of which is low recruitment given that less potential participants are present at the cancer center due to the COVID-19 pandemic. To reach more potential participants, we will advertise our study by posting advertisement flyers on social platforms such as Facebook, Hospital blogs and University websites. Furthermore, three research assistants will take turn stationing at the cancer center five days per week during working hours to recruit participants.

Attrition can be an additional challenge for this RCT provided that the intervention period will last eight weeks. Cancer survivors may discontinue the study due to physical deterioration and other reasons. To overcome this matter, we will offer an honorarium for study participants in an attempt to minimize the drop-out. Furthermore, we will utilize a mobile social platform to communicate with the participants and to send weekly reminders to attend the upcoming intervention session.

## Conclusion

The Digital-MindCAN program is different from existing mindfulness interventions in four aspects. First, the contents cover issues tailored for cancer survivors including cancer-related stressors (such as permanent surgery scars, physical symptoms, and side effects of medications) thoughts (recurrent thoughts of cancer experiences), emotions (such as fear of cancer recurrence, depression and anxiety), and relationships (such as interactions with caregivers and healthcare professionals). Secondly, the use of videotelephony software (such as zoom) will help overcome such issues as manpower, accessibility of the interventions, distance, available time slots, physical limitations and fear of contracting corona virus during the intervention sessions. Furthermore, the videotelephony system will provide a platform to present Power Point slides, education booklet, mindfulness practice videos, websites and other online resources during the intervention sessions. Thirdly, the real-time group sessions allow the facilitator to deliver interactive patient education; and encourage discussions/support among group members. The combination between human touch and technology is perceived to be beneficial to cancer survivors. Finally, the utilization of the mobile messaging application will facilitate communications between group members and the researchers outside the intervention session.

## Data Availability

No datasets were generated or analysed during the current study. All relevant data from this study will be made available upon study completion.

https://doi.org/10.1186/ISRCTN10756993

## Author Contributions

Conceptualization, methodology, writing-original draft, Writing-Review & Editing: PKY

Methodology, writing-original draft, Writing-Review & Editing: KH

Methodology, Writing-Review & Editing: WJC, NKEM, and YSSG.

